# Discrete cortico-hippocampal circuit states underlie mood state transitions in depression

**DOI:** 10.64898/2026.10.11.26365174

**Authors:** Immanuel G Elbau, Andrew M Lee, Megan Chang, Emma Anderson, Parsa Nilchian, Lindsay Victoria, Nicola Manfredi, Neil Gallagher, Vasilisa Iatckova, Niki Sabetfakhri, John Alvarez, Natalya Slepneva, Indira Summerville, Jolene Chou, Megan Johnson, Claire Ho, Eric Goldwaser, Justin Rossi, Ankit Khambhati, Anusha Allawalla, Daniela Astudillo Maya, Stefano Moia, Cesar Caballero-Gaudes, Amy Aloysi, James Murrough, Faith M Gunning, Ben Zebley, Jonathan D Power, Nolan Williams, Logan Grosenick, Edward Chang, Kristin K Sellers, Andrew Krystal, Charles J Lynch, Conor Liston

## Abstract

The hippocampus has long been implicated in depression, yet the underlying mechanisms remain incompletely understood. Leveraging precision functional mapping in densely sampled individuals and large cross-sectional cohorts, and validating key results in direct intracranial EEG recordings from the human brain, we show that hippocampal connectivity alternates between two discrete and opposing network states on the order of minutes: one dominated by default mode network (DMN) coupling and another by sensorimotor system (SM) coupling. This dynamic switching is a fundamental property of hippocampal function in both healthy brains and in major depressive disorder (MDD). In MDD, this equilibrium is disrupted, such that the DMN-dominant state is overrepresented. In densely sampled individuals, hippocampus-DMN coupling tracks symptom fluctuations over time and normalizes after electroconvulsive therapy (ECT), which drives switches in hippocampal connectivity away from the DMN-dominant state and toward the SM-dominant state—an effect that was closely linked to a positive treatment response. Direct intracranial recordings in patients with MDD identified analogous fluctuations in hippocampal–DMN theta synchrony, which tracks symptom fluctuations over hours and days. Together, these findings identify a dynamic cortico-hippocampal mechanism underlying depression and indicate that successful neuromodulation treatments such as ECT act in part by restoring a dynamic equilibrium in hippocampal network connectivity.

---

Depression is a leading cause of disability worldwide (*1*) and is increasingly understood as a disorder of large-scale brain circuit dysfunction (*2–7*). Yet the mechanisms that govern transitions between episodes of depression and normal euthymic mood states—the neural dynamics that control mood over time—remain poorly understood. The hippocampus has long been implicated in depression (*8*), with numerous studies identifying reductions in hippocampal volume and abnormal connectivity in depression;(*9–13*) stress-induced dendritic atrophy and associated disruptions in the regulation of neuroendocrine stress responses (*14–16*); and impairments in synaptic plasticity and neurogenesis that are restored by antidepressants(*17–20*). Still, the mechanisms by which hippocampal dysfunction gives rise to dynamic fluctuations in mood over time are not well defined, especially at the level of neural circuits.

Recent advances in functional MRI (fMRI) and precision functional mapping (PFM) now allow network organization to be characterized in individual human brains with high spatial fidelity and longitudinal depth (*21*–*26*), enabling new approaches to understanding how changes in network function relate to changes in cognition, mood, and behavior over time. PFM quantifies the unique topography of each individual’s cortical and subcortical networks, revealing functionally meaningful individual idiosyncrasies that are obscured in group-averaged parcellations and analyses. When combined with dense longitudinal sampling—acquiring dozens of scans from the same individual across months or years—this approach has enabled pioneering work linking network topography and fluctuations in functional connectivity (FC) to changes in clinical states and behaviors over time (*27–31*).

Similar methods have already revealed previously unrecognized complexity in hippocampal functional organization (*32*, *33*). However, prior studies were designed to measure static affiliations with cortical networks, not dynamic changes over time. Interestingly, intracranial recordings in rodents have demonstrated state-dependent shifts in hippocampal–prefrontal coupling, underscoring how hippocampal network affiliations may not be fixed but may instead reorganize across behavioral states (*34*). PFM, coupled with dense sampling, now permits the critical question of whether the human hippocampus dynamically shifts its network embedding over time. Addressing this question through dynamic functional connectivity analyses that rule out various confounds is challenging in conventional datasets (*35*). Such analyses require large volumes of high-fidelity fMRI data within individual subjects, new denoising strategies, and rigorous statistical testing against realistic null models to overcome challenges posed by sampling error, physiological artifacts, and overfitting (*35*). In this report, we sought to apply these methods in rich, densely sampled fMRI data to test whether human hippocampal network states reconfigure over time, how they relate to real-world mood state transitions, and whether they are modifiable by therapeutic neuromodulation interventions.

Electroconvulsive therapy (ECT) provides a powerful experimental model for studying these transitions. As one of the most potent and clinically effective antidepressant treatments (*36–39*), ECT induces large, predictable shifts in mood and cognition over days to weeks that gradually dissipate over time, especially in patients undergoing maintenance ECT. These rapid state changes offer a unique opportunity to examine the neural mechanisms by which therapeutic interventions modulate hippocampal–cortical connectivity. Unlike pharmacologic or psychotherapeutic treatments that unfold slowly, ECT’s temporal precision allows repeated within-subject sampling of neural dynamics across multiple cycles of response and relapse.

Here, we first applied this precision approach to a densely sampled individual with major depressive disorder (MDD) undergoing ECT, enabling direct observation of hippocampal circuit-level changes accompanying repeated mood state transitions. We discovered that hippocampal functional connectivity alternates between two discrete and opposing configurations: one coupled to the DMN and another to sensorimotor (SM) systems. Analyses of densely sampled healthy individuals and large cross-sectional datasets confirmed that this dynamic switching is a fundamental property of hippocampal function. Building on this discovery, we demonstrate that the balance between hippocampal states is disrupted in MDD, with dynamic fluctuations in hippocampal network state occupancy tracking symptom severity, and that this imbalance normalizes with effective therapeutic neuromodulation. Finally, we used direct intracranial recordings of hippocampal network function in depressed patients to validate this mechanism and evaluate its neurophysiological substrates, identifying analogous fluctuations in hippocampal–DMN theta synchrony that track symptom fluctuations over hours and days. Together, these convergent findings identify a dynamic cortico-hippocampal mechanism underlying depressive mood states switches and provide a unifying framework linking circuit dynamics, mood regulation, and treatment response.

## Results

### Rapid-acting neuromodulation triggers a shift in hippocampal connectivity

To characterize circuit-level connectivity changes accompanying rapid mood state shifts induced by ECT, we acquired repeated multi-echo fMRI scans in an individual receiving maintenance ECT for MDD, yielding a densely sampled serial imaging dataset comprising 39 high-fidelity fMRI scans over six months spanning 16 ECT sessions (**Fig. 1a**). As expected, ECT produced rapid but transient symptom improvements (**Fig. 1a**, lower panel). Depressive symptoms re-emerged within weeks after each course of ECT, such that the patient’s depression severity was tightly correlated (Spearman ρ=0.73, p<10^-6^) with the time elapsed since the last treatment—a metric we term ‘ECT-Distance’ (**Fig. 1b**).

**Figure 1.**
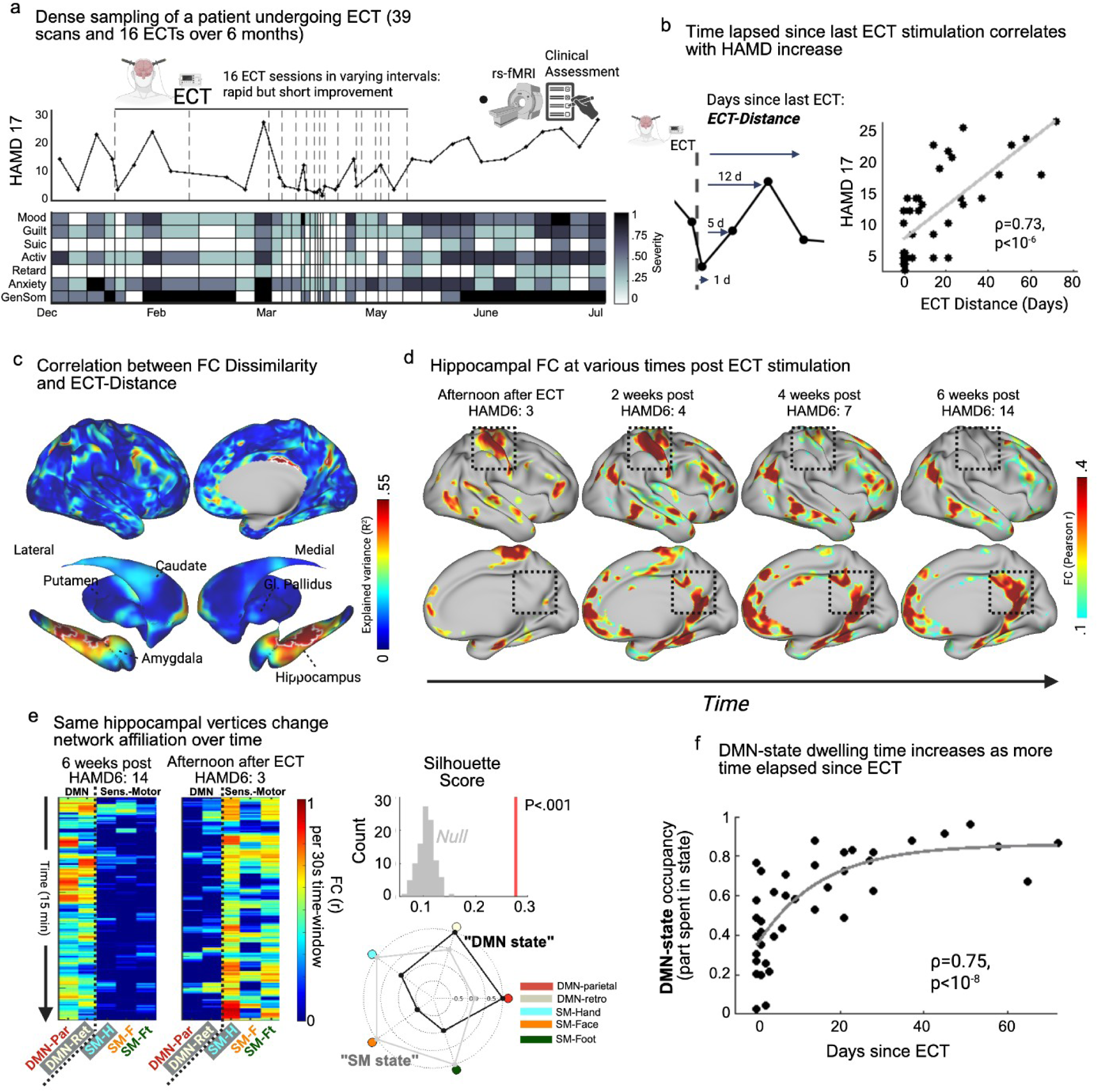
Rapid-acting neuromodulation triggers a shift in hippocampal connectivity states in a densely sampled patient. (a) Longitudinal course for one patient with MDD scanned 39 times while receiving 16 ECT sessions over ∼6 months. Top: HAMD-17 scores show repeated improvement after ECT and gradual relapse. Bottom: item-level heat map (columns = visits) illustrates symptom components acquired at each assessment. (b) “ECT-Distance”—days since the most recent treatment (cartoon, left)—tracked depressive severity across visits (right; Spearman ρ=0.73, p<10⁻⁶). (c) Brain-wide regression of session-wise FC dissimilarity against ECT-Distance (permutation- and multiple-comparisons–corrected) identified the anterior–mid hippocampus functional connectivity as most strongly explained by the clinical time course (lateral/medial and subcortical views; white outlines mark FDE corrected significant clusters). (d) Hippocampal whole-brain FC at representative post-ECT intervals shows a gradual transition from a somatomotor-coupled pattern immediately after treatment to a default-mode–coupled pattern weeks later (boxes highlight hippocampal territory). (e) Time-resolved analysis of the same hippocampal vertices (30-s sliding windows) illustrates shift in affiliation with SM and DMN networks in scans acquired at the afternoon after ECT vs. 6 weeks post ECT. At intermediate time points, both network affiliations recur within a single scan, reflecting alternating hippocampal connectivity states (see panel f). k-means clustering (k=2) yielded two recurrent centroids (“SM state” and “DMN state”), with stability exceeding a phase-shifted null (silhouette score ≈0.3, p<0.001; centroid radar plot for most distinctive networks at right). (f) DMN-state occupancy (fraction of windows per scan assigned to the DMN-dominant centroid) increased with ECT-Distance (Spearman ρ=0.75, p<10⁻⁶; curve = nonparametric fit), linking the return of depressive symptoms to reinstatement of the DMN-dominant hippocampal state.

To identify neural substrates most closely linked to symptomatic responses to ECT, we first conducted an unbiased, whole-brain search for neuroanatomical areas exhibiting the most pronounced shifts in functional connectivity (FC) in association with time elapsed since the last ECT treatment. For each brain vertex, we computed its whole-brain FC profile separately for each of the 39 fMRI scanning sessions. We then quantified session-to-session connectivity reconfigurations for a given vertex by measuring the dissimilarity of its FC profile over time (operationalized as 1 – r^2^, where r = the correlation between its FC profile in a given session and its profiles across other sessions), with larger dissimilarity scores indicating larger FC reconfigurations. Finally, to identify brain areas exhibiting large reconfigurations that accumulate with time elapsed since the last ECT treatment, we regressed dissimilarity scores for each session on ECT-distance. The bilateral hippocampus emerged from this analysis as the single region exhibiting connectivity reconfigurations that were most strongly linked to ECT treatments (**Fig. 1c**; **Supplementary Fig. S1**), with secondary effects observed in the ventral striatum and posterior cingulate.

These effects were most pronounced in the anterior and middle hippocampus (**Fig. 1c**). Immediately following ECT, when symptoms were lowest, it was almost exclusively coupled to the sensorimotor (SM) system ( **Fig. 1d**). As time passed and symptoms returned, connectivity gradually transitioned through a period of shared coupling with both the SM and default mode network (DMN) systems (**Fig. 1d,** 2 and 4 weeks post-ECT), eventually returning to a state of almost exclusive coupling with the DMN (**Fig. 1d**, 6 weeks post-ECT). Although prior literature implies complex topology of hippocampal network affiliation (*40*), the pronounced and sustained shift into an almost exclusively SM-affiliated state—rather than the canonically assumed DMN coupling—was unexpected. Furthermore, the hybrid connectivity pattern observed 2 and 4 weeks post-ECT raised the question as to whether hippocampal connectivity might alternate between affiliations with the DMN and SM on shorter time scales.

**Figure 2.**
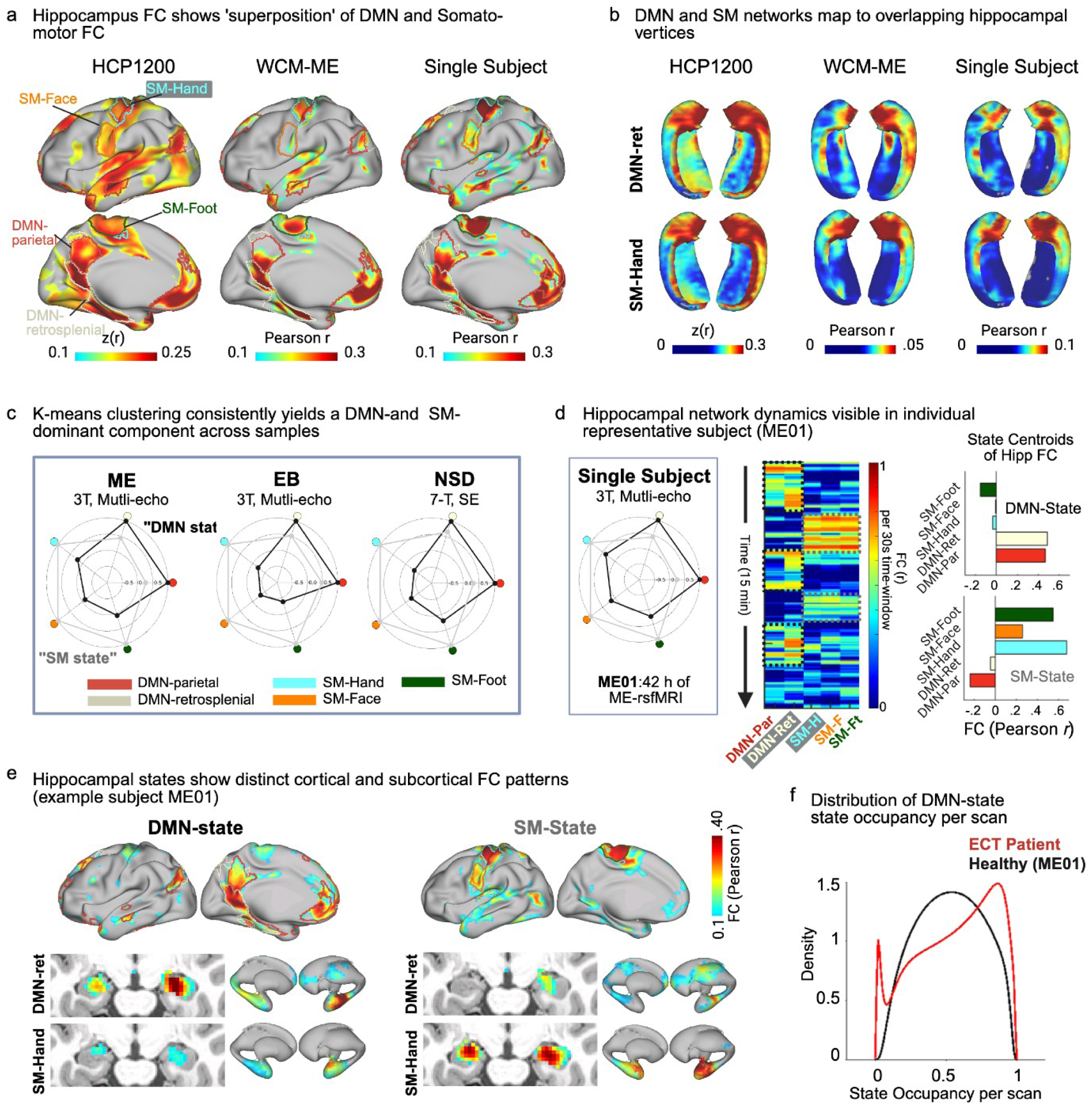
Dynamic switching between DMN- and SM-dominant hippocampal states is a fundamental principle of hippocampal function. (a) Time-averaged hippocampal FC maps from a population cohort (HCP1200), dense-sampled multi-echo fMRI cohort (WCM-ME), and a representative single subject (ME01) show apparent “superposition” of DMN and SM connectivity. Fisher z–transformed correlation values are shown for HCP1200, and Pearson correlation coefficients are shown for WCM-ME and single-subject data. (b) Projecting average DMN-retrosplenial and SM-Hand network connectivity onto hippocampal surfaces reveals spatial overlap of both network’s FC along the anterior CA1–subiculum boundary across datasets (Dice=0.47±0.05; p<0.001 vs. spatial null). (c) Time-resolved k-means clustering of hippocampal-to-cortex FC robustly separated epochs into DMN- and SM-dominant states across three independent dense-sampling cohorts (ME, EB, NSD). (d) In a representative subject (ME01, >40h fMRI), the hippocampus alternated between states on the timescales of one to two minutes, with distinct temporal blocks corresponding to SM and DMN dominance. Right panels: cluster centroids show comparable strength of hippocampal coupling across dominant networks. (e) Whole-brain FC patterns differed markedly between states, with DMN-state showing preferential coupling to retrosplenial and medial-parietal regions, and SM-state to motor and premotor territories; subcortical coupling changes were specific to the hippocampus (see also Supplementary Fig. S3. (f) State occupancy distributions were unimodal in a healthy subject but bimodal in the ECT patient, illustrating stable state balance in health versus pathological DMN bias in depression.

Dynamic functional connectivity analyses are widely used to capture time-varying reconfigurations of large-scale brain networks that are not evident in static, scan-averaged measures, and to identify recurring connectivity states that reflect shifts in functional integration over time(*41–44*). By estimating functional connectivity within temporally localized windows, this approach enables tracking of how a given brain region transiently changes its network embedding during ongoing brain activity. To test whether DMN- and SM-dominant hippocampal connectivity states were also detectable within single scanning sessions, we performed a time-resolved analysis in which connectivity between the hippocampus and 19 cortical networks was estimated within sliding windows across scans, followed by k-means clustering to identify discrete brain states. This approach allowed us to test whether a the hippocampus dynamically altered its network allegiance over time on the order of minutes ( **Fig. 1e**). Statistical rigor was ensured by testing state solutions against phase-shifted surrogate data, evaluating the optimal number of states (i.e. the state solution maximizing the silhouette score), and evaluating stability across a range of parameters (**Supplementary Fig. S2**). This analysis revealed two reproducible states, one dominated by DMN coupling and the other by SM coupling (i.e. a 2-cluster solution maximized the silhouette score, with statistically significant clustering as compared to a phase-shifted null distribution, p<0.001; **Fig. 1e**; for additional data on robustness and reproducibility of two distinct network states, see **Supplementary Fig. S2**). In scans immediately following ECT stimulations, occupancy of the DMN-dominant state was markedly reduced, then gradually increased as time elapsed, closely paralleling both ECT-Distance (Spearman ρ=0.75, p<10⁻⁶; **Fig. 1f**) and symptom recurrence (Spearman ρ=0.61, p=3×10⁻⁵).

### Dynamic switching between DMN and SM states is a fundamental principle of hippocampal function

To determine whether these dynamics reflect a general principle of hippocampal functioning in other MDD cases and in the healthy brain, we first examined time-averaged hippocampal connectivity in healthy individuals, leveraging both a large-scale population cohort (HCP1200; n = 1,206) and multiple densely sampled high-fidelity fMRI datasets (WCM-ME: n = 5 subjects, with 322 ± 312 minutes of scan time per subject; Ekskalibur (EB): n = 10 subjects, with 407 ± 18 minutes / subject; Natural Scenes Dataset (NSD): n = 8 subjects, with 115 ± 36 minutes / subject; for details, see **Supplementary Table S1**). We found that in conventional functional connectivity analyses averaging across each subject’s entire time series (up to 47 hours of fMRI), the anterior-to-mid hippocampus consistently showed stable functional connections with two well-defined brain networks (**Fig. 2a**)—the default mode network (DMN) and the somatomotor (SM) network—which are typically spatially independent and uncorrelated (mean DMN–SM FC in WCM-ME: r = –0.05 ± 0.03). In line with prior reports of an anterior–posterior gradient (*32*), the posterior hippocampus coupled primarily to the medial parietal network (**Supplementary Fig. S3**). We hypothesized that this co-connectivity of the anterior–mid hippocampus with two independent networks—apparent in several prior studies (*27*, *32*, *33*)—may reflect a spatial "superposition" of two separate states of hippocampal-cortical coupling that alternate over time.

To begin to test this, we projected DMN- and SM-network FC onto the hippocampal surface, by correlating average DMN or average SM network BOLD signal time series with voxelwise hippocampal time series. This revealed highly similar patterns of hippocampal connectivity with both networks across cohorts, and clear spatial overlap of both networks’ FC along the anterior CA1–subiculum boundary (mean Dice = 0.47 ± 0.05; **Fig. 2b**). This effect was significant compared to a spatially preserved null, confirming the overlap was not due to random chance or simple anatomical proximity (p < 0.001 in the majority of subjects) across thresholds up to the 90th percentile.

To confirm whether this apparent spatial superposition arises from a dynamic process in which hippocampal connectivity spontaneously alternates between two distinct, recurring states, we performed a time-resolved connectivity analysis, as in Fig. 1e. In this approach, hippocampal–cortical functional connectivity was computed within short sliding windows (30 seconds) across each scan, allowing us to track second-to-minute scale fluctuations in network affiliation. This analysis was made possible by the availability of three rich, deeply sampled datasets—multiband multi-echo acquisitions optimized for medial temporal lobe signal (WCM-ME, EB) and ultra–high field 7T data (NSD)—with 1.6-12.4 hours of fMRI data collected per subject. This depth of sampling enabled us to test for the periodic recurrence of distinct connectivity configurations across extended time spans, rather than being limited to inferences from the short (∼5-10 min) datasets typical of most fMRI datasets, using the methods described for Fig. 1e to ensure statistical rigor (**Supplementary Fig. S4**).

Across all three densely sampled healthy control cohorts, this analysis consistently separated temporal epochs into a DMN-dominant state and an SM-dominant state (**Fig. 2c**). These dynamics were clearly observable at the individual level, where a representative healthy subject’s hippocampus alternated between both states several times within a time scale of 15 minutes (**Fig. 2d**). The strength of hippocampal coupling to the dominant network was comparable across the two states (**Fig. 2d**, right panel), indicating that the SM state was not simply a loss of DMN connectivity with residual noise, but a robust and equally expressed configuration characterized by strong hippocampal coupling to sensorimotor systems. We further confirmed that the SM state does not reflect periods of movement: head motion was comparable across states, and the hippocampus did not coactivate with motor activity in either a large-scale meta-analysis or a densely sampled individual (**Supplementary Fig. S5**).

To further characterize these two hippocampal connectivity configurations, we generated whole-brain maps of hippocampal FC in the DMN state and in the SM state in individual subjects, by calculating average hippocampal FC across each state. This analysis confirmed markedly distinct network configurations, with the DMN-state showing preferential hippocampal coupling to retrosplenial and medial-parietal regions, while in the SM-state, the HC was preferentially coupled to motor and premotor territories (**Fig. 2e**). Notably, when projecting DMN- and SM-network connectivity across the subcortex, state-dependent changes were highly specific to the hippocampus; connectivity shifts were observed with the hippocampus but not with other subcortical structures, indicating the absence of global changes in FC driving these states (**Fig. 2e**). This specificity was further evidenced by comparing hippocampal FC dissimilarity across both states to FC dissimilarity of other subcortical structures, which was markedly lower across all samples (**Supplementary Fig. S4**)

To begin to understand whether the occurrence of these two hippocampal network states differed in the patient receiving ECT for MDD, we compared the distribution of DMN-state occupancy across repeated scans in this individual with a representative healthy subject. Over many scans, the healthy control subject’s DMN-state occupancy formed a stable, unimodal distribution centered around a state occupancy of 0.6, indicating a consistent equilibrium between DMN and SM states in which the DMN state is expressed around 60% of the time. In stark contrast, the ECT patient showed a bimodal distribution, underscoring fluctuations between extremes of DMN or SM dominance (**Fig. 2f**).

Together, these observations from thousands of patients, and hundreds of hours of fMRI data in individual subjects indicate that dynamic hippocampal connectivity reconfigurations between two distinct states involving the SM and DMN networks are an intrinsic and generalizable feature of hippocampal physiology. In the individual receiving maintenance ECT for MDD, the equilibrium between these two states is disrupted and strongly modified by ECT sessions. This observation motivated us to test whether this imbalance is a core feature of depression and evaluate the extent to which it generalizes to other MDD patients.

### Hippocampal network state imbalance is a generalizable feature of depression

Building on this observation, we next asked whether this imbalance between hippocampal FC states generalizes beyond the single ECT case analyzed in **Fig. 1**. To this end, we first tested whether other individuals with MDD exhibit a consistent bias toward the DMN-dominant state as observed in **Fig. 1**. Using identical multiband, multi-echo (MB–ME) fMRI sequences, we compared hippocampal DMN-versus SM–state occupancy between 23 healthy controls and 124 patients with MDD, the latter scanned while actively depressed with ≥28 min of data per participant. DMN-state occupancy was significantly higher in MDD patients (67%) than in healthy controls (56%; p = 1.4 × 10⁻⁴; **Fig. 3a**), confirming a disease-related shift toward the DMN-dominant state.

**Figure 3.**
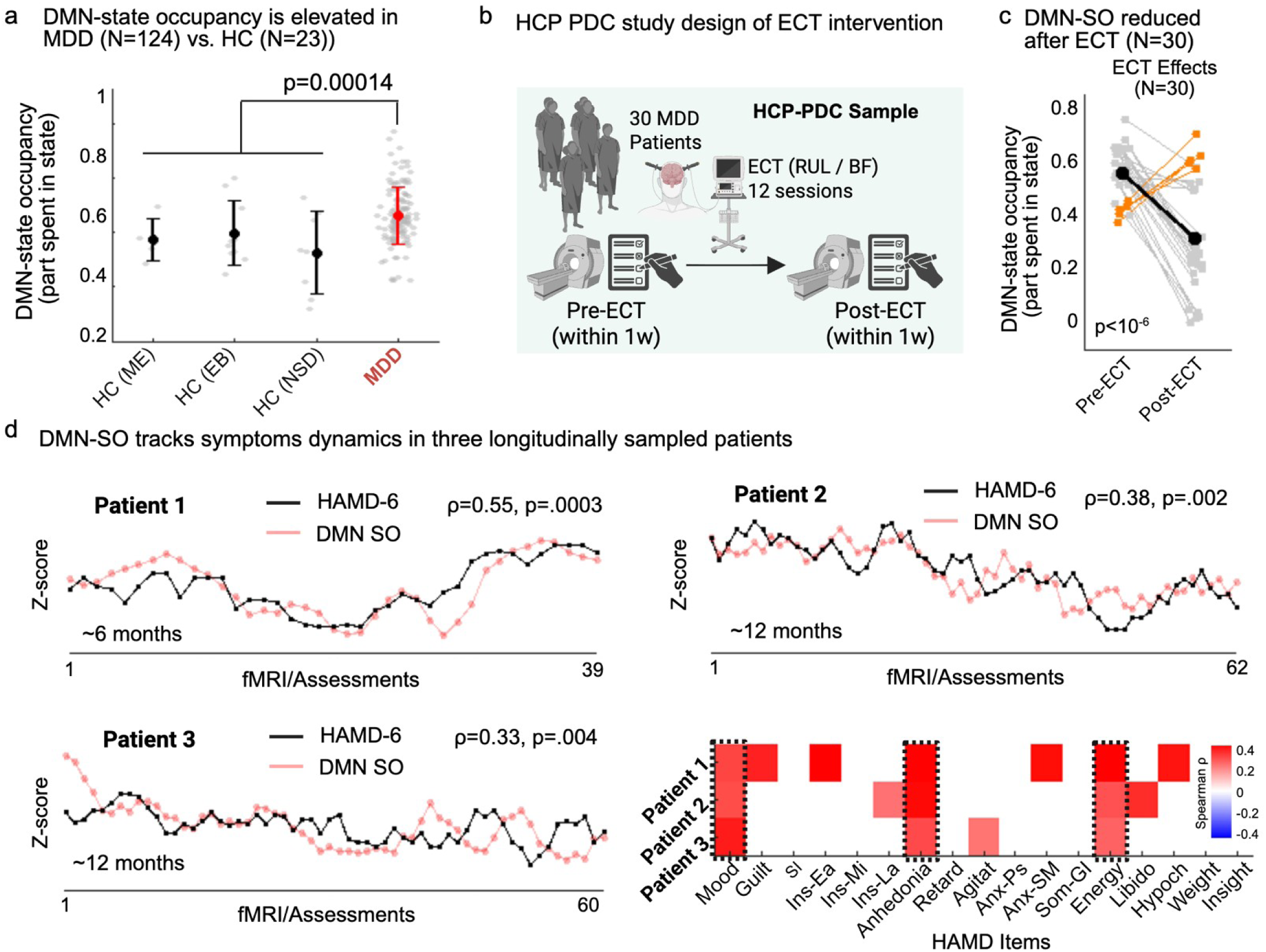
Hippocampal network state imbalance is a generalizable feature of depression. (a) In an independent cross-sectional sample, DMN-state occupancy was elevated in patients with MDD (n=124) relative to three densely sampled healthy control samples (WCM-ME, EB, NSD; n=23; p=0.00014). (b) Schematic of the HCP-PDC ECT cohort (n = 30). c) In this sample, DMN-state occupancy decreased markedly following ECT (*p* < 10⁻⁵). Orange trajectories indicate the four individuals who did not show a post-treatment decrease in DMN-state occupancy; these non-responders exhibited significantly smaller symptom improvements (mean HAMD-6 reduction ≈7 points below group average). (d) In three densely sampled MDD patients (39–62 scans across ∼6–12 months), fluctuations in DMN-state occupancy tracked depressive symptom severity (HAMD-6; Spearman’s ρ=0.55, 0.38, 0.33; all p<0.005). Right panel: correlations were strongest for core symptoms of mood, anhedonia, and low energy. WCM-ME: WCM Multi Echo data set; EB: Ekskalibur data set; NSD: Natural Scenes Data Set.

Having established that MDD is characterized by elevated occupancy of the DMN-dominant hippocampal state, we next sought to test whether ECT rescues this imbalance in an independent cohort (the HCP-PDC ECT dataset; for details, see **Supplemental Table S1** and (*45*)), comprising 30 patients with treatment-resistant depression who received fMRI scans before and after a standard 3–4 week course of right-unilateral or bitemporal ECT. Consistent with the effects observed longitudinally in Fig. 1, ECT drove a shift in hippocampal connectivity toward the SM-dominant state (p < 10⁻⁵; **Fig. 3b-c**). Of note, these shifts were most pronounced in ECT responders. These results confirm that effective therapeutic neuromodulation restores the balance between DMN- and SM-dominant hippocampal states.

Finally, we asked whether hippocampal state dynamics track spontaneous symptom fluctuations outside of structured treatment. To address this, we analyzed three densely sampled patients—each monitored longitudinally for up to one year with repeated fMRI sessions (up to 62 sessions) and concurrent clinical assessments, including the ECT patient from Fig. 1. In all three individuals, week-to-week changes in DMN-state occupancy closely tracked changes in depression severity measured by the HAMD-6 (Spearman’s ρ = 0.55, 0.38, and 0.33; all p < 0.005; **Fig. 3d**; see **Supplementary Fig. S2d, i, n** for results across a range of clustering parameters), with the strongest associations observed for core symptoms of depressed mood, anhedonia, and low energy (**Fig. 3d**, lower right). These results demonstrate that hippocampal state imbalance not only normalizes with treatment but also fluctuates in tandem with the natural ebb and flow of depressive symptoms on the order of weeks, underscoring its role as a dynamic marker of depression severity.

### Direct intracranial recordings link hippocampal-DMN synchrony to depression severity

While the fMRI BOLD signal provides an indirect measure of neuronal activity filtered through the hemodynamic response, intracranial EEG (iEEG) enables direct assessment of neural synchrony and network activity in the human brain. To test whether the fMRI-defined hippocampal state imbalance is also detectable in direct recordings of neuronal circuit dynamics in the human brain, we analyzed stereo-EEG recordings from three patients with treatment-resistant depression enrolled in a closed-loop deep brain stimulation (DBS) trial (*46*) (NCT04004169, 10 bilateral depth leads; ∼160 contacts per subject). Each patient underwent a 10-day inpatient admission with continuous iEEG recording and up to 74 serial HAMD-6 assessments. Electrode contacts were functionally mapped using pre-implantation fMRI. Our initial analyses focused on ipsilateral hippocampal–DMN coupling, probed through DMN contacts that were consistently sampled across all individuals. To quantify hippocampal–DMN coupling, we computed theta-band (4–8 Hz) phase-locking value (PLV) between electrode pairs. Theta-phase synchrony provides a direct electrophysiological index of long-range communication between the hippocampus and prefrontal cortex, and has been implicated in internally oriented cognition, memory retrieval, and affective regulation(*47–49*).

Across all three patients, theta-band PLV between the right hippocampus and the DMN consistently tracked within-person fluctuations in depression severity (Patient 1: ρ=0.39, p=0.0015; Patient 2: ρ=0.47, p=9×10⁻⁵; Patient 3: ρ=0.31, p=0.006; **Fig. 4a,c,e**). We confirmed post-hoc that correlations were consistently highest in the theta-band (rank 1 of 5) vs. neighboring frequency bands (delta to gamma) across all three patients. The effect was strongest for DMN electrode contacts, compared to contacts in other areas (**Fig. 4b,d,f**). It was also lateralized to the right hippocampus (**Fig. 4b,d,f**), with weaker correlations for the left hippocampus, consistent with prior findings of right-biased hippocampal–DMN engagement in depression (*50*, *51*). Comparable results emerged when averaging hippocampal theta-PLV across all ipsilateral DMN nodes (**Supplementary Fig. S6**). Because electrode coverage did not include sufficient sampling of cortical somatomotor territories, we could not directly assess hippocampal–SM coupling or state switching as observed in the fMRI analyses. Together, these analyses of intracranial recording data provide direct neuronal evidence that hippocampal–DMN theta synchrony dynamically tracks depressive symptom expression in vivo. Moreover, because these continuous recordings captured up to 74 repeated mood assessments within 10 days, they demonstrate that hippocampal–DMN coupling tracks mood expression on the hour-to-hour timescale, extending the fMRI findings from week-to-week symptom dynamics in Figs. 1 and 3 to momentary mood regulation in Fig. 4. This convergence across temporal resolutions—from slow hemodynamic changes to rapid electrophysiological synchrony—supports a central role of hippocampal–DMN coupling in the fine-grained modulation of affective state.

**Figure 4.**
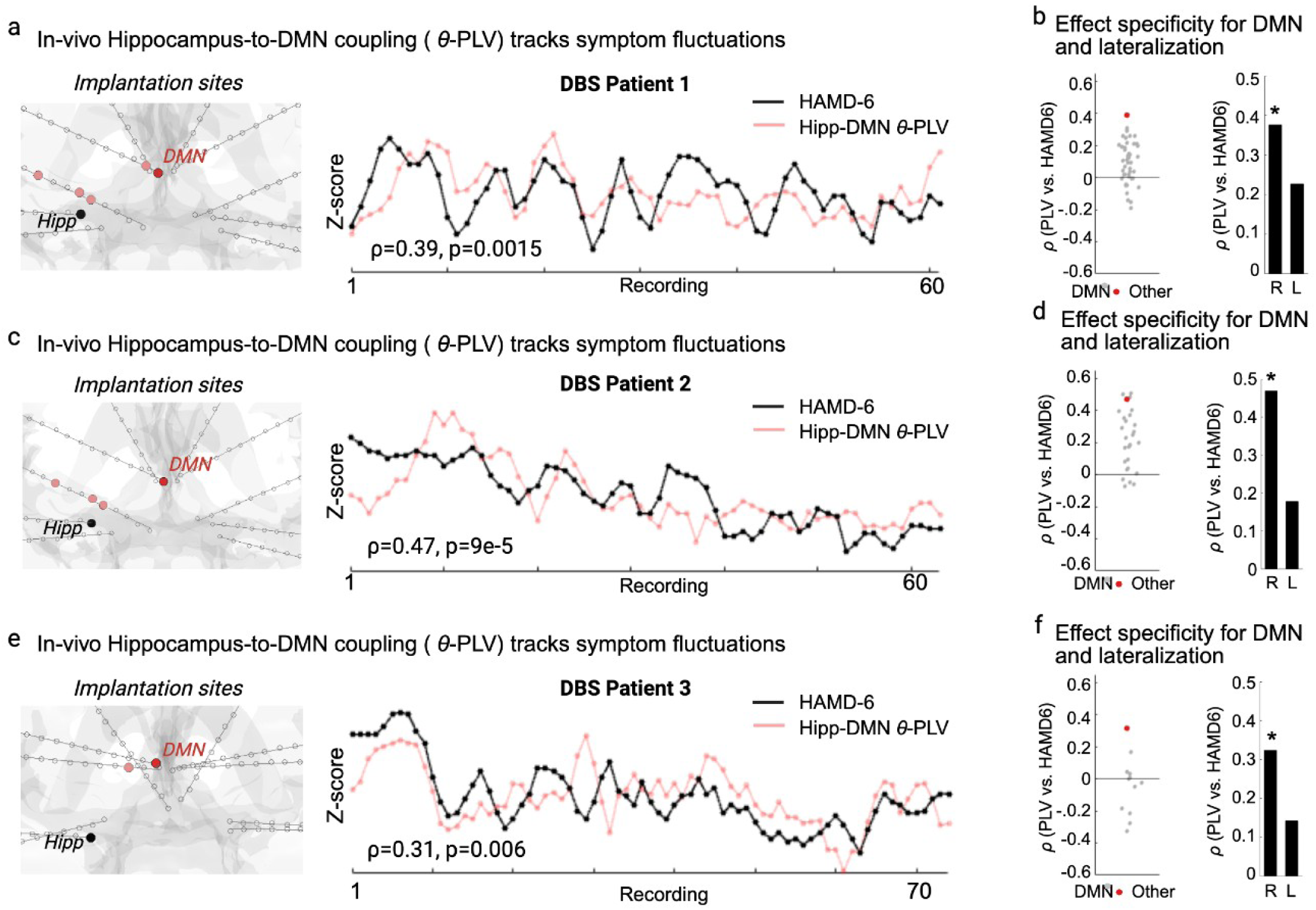
Direct intracranial recordings link hippocampal-DMN synchrony to depression severity. Data are from three patients with treatment-resistant depression implanted with bilateral depth electrodes (∼160 contacts total) and monitored for 10 days with continuous intracranial EEG and up to 72 serial HAMD-6 assessments. Electrode contacts were assigned to precision functional-mapping–derived networks using pre-implantation multi-echo fMRI. (a, c, e) In-vivo hippocampus-to-DMN coupling, computed here with respect to electrode contacts in cortical DMN sites that were consistently sampled across all three subjects. (For hippocampus-to-DMN coupling in traces averaged across all DMN sites in each subject, see Fig. S6, with similar results.) *Left:* Implantation sites show the hippocampus (black) and an electrode contact in a cortical DMN site sampled in all three subjects (red). Pink dots depict DMN contacts that were not consistently sampled. . *Right:* z-scored trajectories of depressive symptoms (HAMD-6, black) and hippocampus–DMN phase-locking value (PLV, red) in the theta band (4–8 Hz). Each patient shows a significant within-subject correlation between HAMD-6 and hippocampus–DMN θ-PLV (Spearman’s ρ and p values indicated). (b, d, f) *Left*: correlation between hippocampus–DMN θ-PLV (red) and depressive symptom severity, contrasted against PLV correlations for the hippocampus with all other cortical contacts (gray). *Right*: correlation between DMN θ-PLV and HAMD-6 for right versus left hemisphere (R, L; * indicates significance). Across all patients, correlations were specific to DMN contacts and right-lateralized, confirming the spatial specificity of hippocampal–DMN synchrony in tracking depressive symptom fluctuations.

## Discussion

Our findings identify a disruption in the dynamic balance between two opposing hippocampal–cortical network states as a core circuit mechanism in depression. Across longitudinal, cross-sectional, and electrophysiological data, we show that hippocampal–DMN hyperconnectivity—a widely studied feature of MDD (*52*)—reflects not a fixed abnormality, but a failure of dynamic regulation: the hippocampus becomes trapped in its DMN-coupled state and fails to re-engage its sensorimotor (SM)-coupled configuration. This breakdown in state flexibility emerged consistently across data types, analytic methods, and patient cohorts and was validated directly by intracranial recordings demonstrating that hippocampal–DMN theta synchrony dynamically tracks depressive symptom fluctuations. ECT acts in part by rescuing this disruption, restoring the SM-coupled configuration and mitigating the dominance of the DMN-coupled state. Together, our results identify a dynamic, mechanistically grounded, and physiologically validated circuit framework for understanding how hippocampal network dysfunction contributes to mood pathology and recovery.

The pattern we describe—a “superposition” of hippocampal connectivity with both default mode (DMN) and sensorimotor (SM) networks—may also explain potentially conflicting and otherwise puzzling observations reported elsewhere in the literature. Although not extensively discussed, other studies mapping hippocampal connectivity gradients or connectivity differences across hippocampal subdivisions have shown that anterior and mid-hippocampal regions show robust coupling not only to DMN territories but also to motor and premotor cortices, despite the fact that the DMN and SM networks are otherwise independent and spatially nonoverlapping. This pattern has been observed across methodological approaches, including precision mapping resting-state and pharmacological fMRI (*27*, *32*, *33*), ultra–high-field imaging (*53*), and task-fMRI (*54*). Yet this convergence of DMN and SM affiliations within the same hippocampal territory is difficult to reconcile with models assuming static network membership or topographical connectivity gradients. The most parsimonious explanation—and the one supported by our data—is that these apparent overlaps arise from alternating hippocampal network affiliations that fluctuate over time between DMN- and SM-dominant states. Our observation that DMN state occupancy rates also fluctuate over time within individuals in response to treatment and with spontaneous changes in mood may also explain in part why some studies have reported abnormally increased connectivity within the DMN in depression(*52*), while others have observed the opposite (*55*), or a combination of the two (*56*). Our results suggest that heterogeneity in sampling with respect to treatment and disease state could contribute to discrepant DMN connectivity findings.

### A Framework for Depression as a Dynamic State Imbalance

This network state imbalance that we observed here may reflect a fundamental trade-off between two opposing modes of cognition. We propose that the hippocampus is central to orchestrating the dynamic switching between an internally-focused brain state supporting self-referential thought (mediated by DMN coupling) and a prospective, action-oriented state for engaging with the world (mediated by SM coupling). In the healthy brain, these network states alternate fluidly according to cognitive demands. In contrast, depression may be understood in part as a breakdown of this dynamic equilibrium, in which the system becomes pathologically biased toward the internally-focused DMN state, resulting in a persistent, self-referential trap.

The link between hippocampus–DMN coupling and internal cognition is well-established. Across human and animal studies, synchrony between the hippocampus and DMN hubs such as the medial prefrontal cortex supports autobiographical memory, episodic prospection, and the planning of internally generated trajectories (*54*, *57*, *58*). This state provides a neural substrate for constructing and maintaining a model of oneself in the world. In contrast, hippocampus–SM coupling may reflect an externally oriented, adaptive mode for engaging with the environment. This state is not simply a reflection of motor output but is instead recruited for prospective simulation, such as envisioning future action sequences and organizing action-outcome maps to guide flexible behavior (*59*, *60*). This aligns with theoretical accounts of the hippocampus binding cortical information into “memories of the future” that imply action (*61*). In this view, hippocampal–DMN hyperconnectivity in depression, which has been observed in numerous studies (*51*, *62*), is not an isolated circuit pathology but rather, one manifestation of a broader systems-level failure: the inability to disengage from the internal, self-referential mode and flexibly switch to the prospective, action-oriented mode needed for adaptive behavior. The therapeutic effect of ECT, which has been shown to increase hippocampus-SM coupling (*63*, *64*), may stem from its ability to restore this dynamic balance.

### Interpretation of N=1 studies

Our use of a deep longitudinal sampling ("N-of-1") approach was critical in this study. In this approach, each individual serves as a complete study, with dense sampling enabling well powered, within-subject statistical inferences. This method, which has a rich history in foundational neuroscience, derives power from extensive within-subject data, with consistency across individuals serving as replication. Our findings from three densely-sampled patients (**Fig. 3d**) represent three independent replications. This logic aligns with a formal inferential framework where a finding’s generality is established by the proportion of individuals who show the effect. By this standard, the 3/3 replication—in both our longitudinal fMRI and our sEEG subjects, and complemented by our analysis of large cross-sectional datasets—provide strong evidence for generalizability, revealing person-specific dynamics often missed in group averages (*65*).

### Limitations

Several limitations should be considered. First, our findings are correlational and do not establish a causal link between hippocampal state occupancy and depressive symptoms. Future studies using targeted circuit manipulation will be needed to determine whether persistent DMN-state occupancy is a cause or a consequence of the depressed state. Second, as noted above, our study relies on a deep longitudinal sampling ("N-of-1") approach that has enabled pioneering new discoveries in multiple contexts(*22*, *27*, *29*, *30*, *66–68*), but further work will be required to understand the degree to which these findings generalize to other individuals, other forms of depression, and whether they associate with specific clinical symptom profiles. Mitigating this concern, in a cross-sectional analysis of 124 patients with MDD, DMN-state occupancy was markedly elevated compared to healthy controls, and completely outside the normal range in ∼20% of MDD patients. Third, our direct electrophysiological validation comes from a small cohort of patients with treatment-resistant depression requiring invasive monitoring. While providing unprecedented neurophysiological data recorded from the living human brain, the generalizability of these specific findings to the broader MDD population warrants further investigation. Finally, while we link the SM-dominant state to prospective cognition based on a wealth of prior literature, its precise functional role is not well understood. Future work using behavioral probes could more directly characterize the cognitive processes associated with these dynamic hippocampal states.

### Conclusion and Future Directions

Our findings establish an imbalance between two dynamic hippocampal–cortical states as a mechanistic signature of depression, identifying both a biomarker and a modifiable circuit target for intervention. By demonstrating that depression involves a failure of the hippocampus to flexibly alternate between DMN- and SM-dominant configurations, this work reframes hippocampal–DMN hyperconnectivity—long observed in depression —not as a static abnormality, but as a manifestation of impaired network switching. The restoration of this equilibrium with effective neuromodulation suggests that interventions such as ECT act by re-engaging an intrinsic dynamic rather than inducing compensatory reorganization. This framework advances current models of depression from a focus on pathological activation toward one centered on the loss and restoration of neural flexibility. It also provides a more precise mechanistic target for next-generation therapies: the hippocampal–cortical circuits that regulate transitions between integrative, DMN-dominant modes and action-oriented, SM-dominant modes of brain function. Future interventions—ranging from network-targeted TMS and focused ultrasound to adaptive closed-loop stimulation—could be designed not merely to suppress pathological patterns but to re-establish the brain’s capacity for adaptive state switching. By defining a core, multimodally validated circuit mechanism that links hippocampal dynamics to mood regulation and recovery, this work lays the foundation for state-based treatments aimed at restoring the brain’s fundamental dynamic equilibrium.

## Data, code, and materials availability

Code used for preprocessing, dynamic connectivity/state analyses, and figure generation will be made publicly available at the time of publication (repository link and DOI to be provided). De-identified derived data products supporting the main figures (e.g., hippocampal state centroids, state-occupancy measures, summary statistics, and analysis-ready outputs) will be deposited in a public repository at the time of publication (accession/DOI to be provided). The Natural Scenes Dataset (NSD), Ekskalibur (EB), and HCP-PDC ECT datasets are publicly available from their respective repositories. The Weill Cornell multi-echo dataset (WCM-ME) and densely sampled clinical imaging dataset(s) are available from the corresponding authors upon reasonable request and completion of required data use agreements/approvals. Data from the ongoing Weill Cornell TMS/iTBS clinical trial (WCM-TMS) will be made available following trial completion and primary reporting, subject to institutional approvals and participant privacy protections.

## Author contributions

I.G.E., C.J.L., and C.L. conceived the study. I.G.E. and C.J.L. designed the fMRI analyses. I.G.E. developed analysis code and performed statistical analyses, in collaboration with L.G., J.D.P., and N.G. I.G.E. generated figures and visualizations. MRI data collection and clinical phenotyping at Weill Cornell were led by I.G.E., C.L., C.J.L., F.M.G., and B.Z., together with M.C., N.M., V.I., I.S., J.C., M.J., C.H., E.G., J.R., P.N., and L.V. UCSF intracranial EEG data collection and analyses were led by A.M.L., E.A., N.Sa., J.A., N.Sl., E.C., K.K.S., A.K., and N.W. A.A. and J.M. contributed to ECT analysis and interpretation of ECT-related results.

S.M. and C.C.-G. contributed the Ekskalibur dataset and provided analytic expertise. C.J.L. and C.L. supervised the project. I.G.E. wrote the initial manuscript draft, and all authors reviewed and edited the manuscript.

## Methods

### Datasets

We analyzed data from several independent sources (**Supplementary Table S1**). Population-scale data were obtained from the Human Connectome Project (HCP1200; n = 1,206; 2,412 scans (*69*)). Densely sampled healthy controls came from the Weill Cornell multi-echo (WCM-ME; n = 5, up to 322 min (*26*)), Ekskalibur (EB; n = 5, up to 407 min), and NSD datasets (n = 8, up to 115 min). Patient samples included three deeply phenotyped patients with MDD or bipolar II disorder (SIMD, up to 1,736 min collected for over a year), and two treatment cohorts (HCP-PDC ECT: n = 30 (*45*), and WCM-TMS n = 124). The UCSF Intracranial stereo-EEG data (UCSF-iEEG) were collected from three MDD patients implanted with multi-electrode arrays for clinical monitoring at UCSF (see details below). All participants provided written informed consent; study protocols were approved by the relevant institutional review boards.

### MRI Acquisition

#### Weill Cornell Medicine ‘Serial Imaging of major depression’ and ‘ME’ datasets

Data were acquired on a Siemens Magnetom Prisma 3 T scanner at the Citigroup Biomedical Imaging Center of Weill Cornell medical campus using a Siemens 32-channel head coil. Two multi-echo, multi-band resting-state fMRI scans were collected using a T2*-weighted echo-planar sequence covering the full brain (TR 1,355 ms; TE1 13.40 ms, TE2 31.11 ms, TE3 48.82 ms, TE4 66.53 ms and TE5 84.24 ms; FOV 216 mm; flip angle 68° (the Ernst angle for grey matter assuming a T1 value of 1,400 ms); 2.4 mm isotropic voxels; 72 slices; AP phase encoding direction; in-plane acceleration factor 2; and multi-band acceleration factor 6) with 640 volumes acquired per scan for a total acquisition time of 14 min and 27 s. Spin echo EPI images with opposite phase encoding directions (AP and PA) but identical geometrical parameters and echo spacing were acquired before each resting-state scan. Multi-echo T1-weighted (TR/TI 2,500/1,000 ms; TE1 1.7 ms, TE2 3.6 ms, TE3 5.5 ms and TE4 7.4 ms; FOV 256 mm; flip angle 8° and 208 sagittal slices with a 0.8 mm slice thickness) and T2-weighted anatomical images (TR 3,200 ms; TE 563 ms; FOV 256; flip angle 8° and 208 sagittal slices with a 0.8 mm slice thickness) were acquired at the end of each session.

#### Weill Cornell rTMS dataset

MRI data were acquired on a Siemens Magnetom Prisma 3 T machine at the Citigroup Biomedical Imaging Center of Weill Cornell medical campus using a Siemens 32-channel head coil as part of an ongoing TMS trial (NCT04041479). Two multi-echo, multi-band resting-state fMRI scans were collected at each study visit using a T2*-weighted echo-planar sequence covering the full brain (TR 1,300 ms; TE1 12.60 ms, TE2 29.51 ms, TE3 46.42 ms and TE4 63.33 ms; FOV 216 mm; flip angle 67° (the Ernst angle for grey matter assuming a T1 value of 1,400 ms); 2.5 mm isotropic voxels; 60 slices; AP phase encoding direction; in-plane acceleration factor 2; and multi-band acceleration factor 4) with 650 volumes acquired per scan for a total acquisition time of 14 min and 5 s. Spin echo EPI images with opposite phase encoding directions (AP and PA) but identical geometrical parameters and echo spacing were acquired before each resting-state scan. Multi-echo T1-weighted (TR/TI 2,500/1,000 ms; TE1 1.7 ms, TE2 3.6 ms, TE3 5.5 ms and TE4 7.4 ms; FOV 256 mm; flip angle 8° and 208 sagittal slices with a 0.8 mm slice thickness) and T2-weighted anatomical images (TR 3,200 ms; TE 563 ms; FOV 256 mm; flip angle 8° and 208 sagittal slices with a 0.8 mm slice thickness) were acquired at the end of each session.

Details of acquisition parameters for the WCM-ME(*26*), Euskalibur (*70*), NSD, and HCP-PDC ECT (*71*) data sets can be found in the respective references.

#### Anatomical preprocessing and cortical surface generation

Anatomical data were preprocessed and cortical surfaces generated using the Human Connectome Project (HCP) PreFreeSurfer, FreeSurfer and PostFreeSurfer pipelines (v.4.3) (*72*).

#### Multi-echo fMRI preprocessing

Preprocessing of multi-echo data minimized spatial interpolation and volumetric smoothing while preserving the alignment of echoes (*26*). The single-band reference (SBR) images (one per echo) for each scan were averaged. The resultant average SBR images were aligned, averaged, co-registered to the ACPC-aligned T1-weighted anatomical image and simultaneously corrected for spatial distortions using FSL topup and epi_reg programs. Freesurfer bbregister algorithm was used to refine this co-registration. For each scan, echoes were combined at each time point and a unique 6 DOF registration (one per volume) to the average SBR image was estimated using FSL MCFLIRT tool, using a four-stage (sinc) optimization. All of these steps (co-registration to the average SBR image, ACPC alignment and correcting for spatial distortions) were concatenated using FSL convertwarp tool and applied as a single spline warp to individual volumes of each echo after correcting for slice time differences using FSL slicetimer program. The functional images underwent a brain extraction using the co-registered brain extracted T1-weighted anatomical image as a mask and corrected for signal intensity inhomogeneities using ANT N4BiasFieldCorrection tool. All denoising was performed on preprocessed, ACPC-aligned images.

#### Multi-echo fMRI denoising

Preprocessed multi-echo data were submitted to multi-echo ICA (ME-ICA103), which is designed to isolate spatially structured T2*-dependent (neurobiological; BOLD-like) and S0-dependent (non-neurobiological; not BOLD-like) signals and implemented using the tedana.py workflow104 (*73*). In short, the preprocessed, ACPC-aligned echoes were first combined according to the average rate of T2* decay at each voxel across all time points by fitting the monoexponential decay, S(t) = S0e−t/T2*. From these T2* values, an optimally combined multi-echo (OC-ME) time series was obtained by combining echoes using a weighted average (WTE = TE × e−TE/T2*). The covariance structure of all voxel time courses was used to identify main signals in the OC-ME time series using principal component and independent component analysis. Components were classified as either T2*-dependent (and retained) or S0-dependent (and discarded), primarily according to their decay properties across echoes. All component classifications were manually reviewed by C.J.L. and revised when necessary following the criteria described in (*74*). Mean grey matter time-series regression was performed to remove spatially diffuse noise (*75*). Temporal masks were generated for censoring high-motion time points using a framewise displacement threshold of 0.3 mm and a backward difference of two TRs (*76*), for an effective sampling rate comparable to historical framewise displacement measurements (approximately 2–4 s). Before the framewise displacement calculation, head realignment parameters were filtered using a stopband Butterworth filter (0.2–0.35 Hz) to attenuate the influence of respiration (*77*) on motion parameters.

#### Single-echo fMRI denoising

The following denoising procedures were applied to the HCP-PDC ECT, and NSD datasets. The NSD dataset was obtained in an already preprocessed (but not yet denoised) format. Preprocessed single-echo data were then submitted to ICA-AROMA (*78*). All component classifications were manually reviewed by study author C.J.L. and revised when necessary following standardized criteria (*74*). Mean grey matter time-series regression was performed to remove spatially diffuse noise. Temporal masks were generated for censoring high-motion time points, as done for the multi-echo fMRI datasets.

#### Surface processing and CIFTI generation of fMRI data

The denoised fMRI time series was mapped to the individual’s fsLR 32k midthickness surfaces with native cortical geometry preserved (using the -ribbon-constrained method), combined into the connectivity informatics technology initiative (CIFTI) format and spatially smoothed with geodesic (for surface data) and Euclidean (for volumetric data) Gaussian kernels (σ = 2.55 mm) using Connectome Workbench command line utilities (*79*). This yielded time courses representative of the entire cortical surface, subcortex (accumbens, amygdala, caudate, hippocampus, pallidum, putamen, thalamus and brainstem) and cerebellum but excluding non-grey matter tissue. Spurious coupling between subcortical voxels and adjacent cortical tissue was mitigated by regressing the average time series of cortical tissue of less than 20 mm in Euclidean space from a subcortical voxel.

#### Hippocampal segmentations

Hippocampal subfields were segmented with HippUnfold (v≥1.3), which combines U-Net–based segmentation with topology-preserving unfolding into intrinsic coordinates (anterior–posterior, proximal–distal, inner–outer) via Laplace’s equation (*80*). Ex-vivo atlas–guided diffeomorphic registration was used to improve cross-subject alignment. HippUnfold outputs mid-thickness surfaces and subfield labels in native and unfolded space. Subject-level analyses were performed in native space; for group analyses (e.g., Fig. 1), label maps were normalized to MNI space using TemplateFlow (*81*) and projected onto the hippocampal surface template.

#### Precision mapping of functional of brain networks in individuals

A detailed description of PFM procedures can be found in (*28*). A functional connectivity matrix summarizing the correlation between the time courses of all cortical vertices and subcortical voxels across all study visits was constructed. Correlations between nodes 10 mm or less apart (geodesic and Euclidean space used for cortico–cortical and subcortical–cortical distance, respectively) were set to zero. Correlations between voxels belonging to subcortical structures were set to zero. Functional connectivity matrices were thresholded in such a way that they retained at least the strongest X% correlations (0.01%, 0.02%, 0.05%, 0.1%, 0.2%, 0.5%, 1%, 2% and 5%) to each vertex and voxel and were used as inputs for the InfoMap community detection algorithm (*82*), one of the most widely used approaches for delineating functional brain networks and their boundaries in individuals (*21*, *22*, *25*, *27*). Free parameters (for example, the number of algorithm repetitions) for the Infomap algorithm were fixed across subjects. The total number of communities identified by Infomap is controlled in part by how many connections are retained in the functional connectivity matrix after thresholding. The optimal scale for further analysis across individuals was defined as the graph threshold producing the best size-weighted average homogeneity relative to the median of the size-weighted average homogeneity calculated from randomly rotated networks, as done in (*83*). Size-weighted average homogeneity was maximized relative to randomly rotated communities at the 0.1% graph density and resulted in 89.13 ± 8.04 communities on average across individuals. Each Infomap community was algorithmically assigned to one of 20 possible functional network identities (Default-Parietal, Default-Anterolateral, Default-Dorsolateral, Default-Retrosplenial, Visual-Lateral, Visual-Stream, Visual-V1, Visual-V5, Frontoparietal, Dorsal Attention, Premotor/Dorsal Attention II, Language, Salience, Cingulo-opercular/Action-mode, Parietal memory, Auditory, Somatomotor-Hand, Somatomotor-Face, Somatomotor-Foot, Auditory or Somato-Cognitive-Action) primarily according their functional connectivity and spatial locations relative to a specified set of priors. All algorithmic assignments were manually reviewed by C.J.L. and manually adjusted in the case of an ambiguous assignment. A temporal network (node × node × study visit) summarizing the correlation between the time courses of all cortical vertices and subcortical voxels across study visits was constructed for each patient. Correlations between nodes less than 10 mm apart (geodesic and Euclidean space used for cortico–cortical and subcortical–cortical distance, respectively) were set to zero. Correlations between voxels belonging to subcortical structures were set to zero. Links between layers were generated automatically using neighborhood flow coupling.

#### Clinical Assessments

Patients across all clinical samples were evaluated with the Hamilton Depression Rating Scale (HDRS-17(*84*)), from which the HAMD-6 subscale was derived for analyses (*85*). Depressive symptoms in the UCSF stereo-EEG cohort were assessed using a visual analogue scale for depression (VAS-D (*86*)). For longitudinal datasets, clinical ratings were obtained during the same sessions as fMRI scanning. Treatment response was defined as the percentage reduction in symptom scores from baseline.

### ECT treatments

#### ECT treatments of patient MDD01

The patient received 16 ECT sessions in total, as outlined in Figure 1a. Treatments were 11 bitemporal (BL) at 20% and 5 right-unilateral (RUL) at 75%. The shift from BL to RUL was made due to cognitive side effects. General anesthesia was methohexital with adjunct ketamine for all sessions. EEG seizure duration averaged 53.0 ± 10.4 s across sessions (mean ± SD), with BL: 55.2 ± 11.4 s and RUL: 48.2 ± 6.6.

#### HCP-PDC ECT cohort

Clinical care followed standard protocols not manipulated by the study: seizure threshold was titrated at the first session, treatment began with right-unilateral (RUL) ECT (pulse width 0.3 ms, 800 mA), and patients were switched to bitemporal (BL) ECT when clinically indicated (pulse width 0.5 ms, 8,000 mA as specified in the manual). Treatments were administered every 2–3 days over ∼3–4 weeks until maximal response or remission was sustained for ≥1 week. Research assessments and MRI were obtained pre-ECT and within one week after the index series.

#### Hippocampal functional connectivity

Fully denoised BOLD time series were extracted bilaterally from the anterior hippocampus (geometric anterior three-fifths along the A–P axis). These time courses were correlated with the average time courses of 19 precision functional mapping (PFM)–derived networks to obtain network-wise hippocampal FC values.

#### Dynamic FC and state detection

Dynamic functional connectivity (dFC) was computed between the bilateral anterior hippocampus (geometric anterior three-fifths along the A–P axis) and 19 precision functional mapping (PFM)–derived cortical networks. Fully denoised BOLD time series were segmented into 30-s sliding windows (step size = 1 TR; range 15–60 s tested for robustness). Within each window, hippocampus-to-network correlations were calculated using Pearson correlation (Fisher-z transformed). Windowed dFC matrices were concatenated and clustered using k-means in MATLAB (correlation distance metric; default Lloyd’s algorithm with 100 random restarts. Optimal clustering solutions were determined using the silhouette score, with statistical significance assessed relative to a phase-shuffled null distribution that preserved feature-wise autocorrelation. For each candidate *k*, we computed the mean silhouette score in the empirical data and compared it to the distribution of silhouette scores obtained from surrogate data. We selected the smallest *k* whose empirical silhouette score exceeded the null by the greatest margin while falling within one standard error of the maximum difference. For longitudinal MDD and BD patients, clustering was performed at the individual level to capture within-subject state dynamics. For cross-sectional and group-level analyses, state centroids were derived from healthy control datasets (WCM-ME, EB, NSD) and applied to patient samples to ensure consistency across cohorts. State stability was confirmed across window lengths (**Supplementary Fig. S4**). For visualization, state-defining features (mean hippocampal FC to each network) were summarized in radar plots.

#### Intracranial EEG participants, Acquisition, and Analysis

Three adults (ages 30 to 34, 2 F) with treatment-resistant major depressive disorder (TRD) were recruited as part of a single-center feasibility trial investigating personalized closed-loop neuromodulation with the NeuroPace Responsive Neurostimulation (RNS) system. Inclusion criteria required DSM-V MDD without psychosis, ≥2 years of current depressive episode, failure of ≥4 adequate antidepressant treatments (including ECT), a baseline MADRS >26, and demonstrable variability in HAMD-6 ratings over a 3-day period. Exclusion criteria included bipolar disorder, active suicidal ideation, substance abuse, neurological disorders, severe medical comorbidities, pregnancy or breastfeeding, and inability to comply with follow-up procedures. Each subject underwent surgical implantation of ten stereoelectroencephalography (sEEG) electrodes (PMT Corporation) targeting orbitofrontal cortex, amygdala, hippocampus, ventral capsule/striatum, and subgenual cingulate cortex. Each electrode contained 16 contacts, yielding ∼160 contacts per participant. Recordings were acquired with a 256-channel Nihon Kohden clinical system, sampled at 10 kHz.

#### Pre-implant fMRI and Electrode Classification

Four sessions of pre-operative rsfMRI (60 to 120 min) were acquired (TR = 800 ms, TE = 30 ms, voxel size 2.4 mm isotropic) and preprocessed using fMRIPrep and XCP-D2. Denoising included regression of motion, WM/CSF, and global signal, band-pass filtering (0.008–0.1 Hz), and motion scrubbing. Individualized cortical network parcellations (17 canonical Yeo networks) were derived using multi-session Bayesian modeling. Electrode contacts were mapped to these individualized networks using a support vector machine (SVM) classifier trained on fMRI functional connectivity (FC) profiles. Contacts were retained if posterior probability of assignment exceeded 0.5. Analyses focused on hippocampal–DMN coupling, with the primary analysis shown in Fig. 4 restricted to DMN contacts consistently sampled across all individuals.

#### Clinical Assessments

During a 10-day inpatient admission, depressive symptoms were assessed up to 74 times per subject using the HAMD-6 and a visual analogue scale for depression (VAS-D). These repeated assessments provided within-subject symptom fluctuations for electrophysiological correlation.

#### iEEG Preprocessing

Continuous recordings were segmented into 6-min epochs immediately preceding each mood rating. Data were down sampled to 512 Hz, band-pass filtered (0.5–240 Hz), and notch filtered at 60 Hz harmonics. Channels outside brain parenchyma or with artifactual activity were excluded. Segments with global amplitude >2 SD from the mean were rejected. Signals were re-referenced using common average referencing (CAR).

#### Phase-Locking Value Analysis

For each epoch, signals were filtered into canonical bands: δ (1–4 Hz), θ (4–8 Hz), α (8–13 Hz), β (13–30 Hz), and low γ (30–70 Hz). Instantaneous phase was extracted using the Hilbert transform, and pairwise phase-locking value (PLV) was computed as:

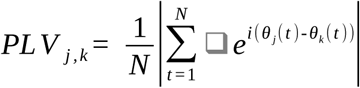

where N is the number of time points and θ*_j_*(*t*) and θ*_k_*(*t*) are the instantaneous phase of signals *j* and *k* at timepoint *t,* and *e^i^* ^(^ *^θ j^* ^(^ *^t^* ^)-*θ*^*^k^* ^(^ *^t^* ^))^ is the complex exponential of the phase difference at each time point. PLVs range from 0 (no synchrony) to 1 (perfect synchrony). Frequency-specific FC matrices were generated for each epoch and aligned with concurrent symptom scores.

#### PLV-behavior associations

PLVs were averaged across hippocampal–cortical contact pairs within each network. Associations between hippocampal–DMN PLV and depression severity were tested using within-subject Spearman correlations. Significance thresholds were determined parametrically. Effects were examined across frequency bands to test for spectral specificity.

#### Statistical Analysis

Unless otherwise noted, statistical tests were two-sided. Exact n, degrees of freedom, and P values are reported in figure legends. Correction for multiple comparisons was performed using Bonferroni or permutation methods.

#### Brain-Wide Dissimilarity Analysis

To identify the brain region whose connectivity dynamics were most robustly explained by the ECT clinical course, we performed a brain-wide multiple regression analysis. For each brain node (vertex or voxel), we first computed its whole-brain functional connectivity (FC) pattern for each of the 39 fMRI sessions. From these, we derived a session-wise similarity vector for each node. The dependent variable for our model was the session-wise dissimilarity, calculated as 1−r2, where r is the similarity value for a given session. A multiple regression model was then fitted at each node, predicting the dissimilarity time-series from two ECT-related predictors: ‘ECT-Distance’ (time elapsed since last treatment) and ‘ECT-Load’ (a cumulative metric). The overall explanatory power of the model at each node was quantified by its cumulative R² value. To identify nodes with a statistically significant R² value, we performed a rigorous permutation test (n=1000). For each permutation, the time-series of the ECT predictors were circularly shifted together to generate a null distribution of R² values while preserving the temporal autocorrelation of the original data. To correct for multiple comparisons across all brain nodes, we accounted for the spatial smoothness of the data. We first calculated Moran’s I, a measure of spatial autocorrelation, to estimate the effective number of independent tests across the brain. A Bonferroni-style correction was then applied using this effective number of tests (α=0.05). The final output was a brain map of R² values, thresholded to show only the nodes that survived this stringent statistical correction.

#### Hippocampal FC Projection and Spatial Overlap

To quantify the spatial overlap of DMN- and SM-related connectivity patterns, we first projected the functional connectivity (FC) map from each network onto the grayordinates of each subject’s hippocampus. Within the hippocampus, each of the two maps was independently thresholded (retaining the top 10% of connectivity values) to create two binary masks, representing the vertices with the strongest connectivity to the DMN and SM networks, respectively. The empirical overlap between these two masks was then calculated using the Dice coefficient: 2×∣A∩B∣/(∣A∣+∣B∣), where A and B are the sets of vertices in each mask. To determine if the observed overlap was greater than expected by chance, we compared it against a spatially constrained null model using a permutation test (n=1000). Our primary null model was designed to preserve the local spatial autocorrelation present in the connectivity maps. To achieve this, the hippocampal grayordinates were first segmented into small, spatially contiguous 3D blocks (e.g., 3×3×3 voxels). For each permutation, these entire blocks were randomly shuffled, and the SM connectivity map was reconstructed based on this new block arrangement. This procedure generates a null map that maintains local smoothness and patchiness but disrupts the global spatial organization. A null Dice coefficient was calculated for each permutation, creating a null distribution. The p-value was then computed as the proportion of null coefficients greater than or equal to the empirically observed Dice coefficient. In cases where 3D coordinate information was unavailable, a fallback 1D block-shuffling procedure along a functionally-defined manifold was used.

## Data Availability

The data supporting the findings of this study may be made available by the corresponding author upon reasonable request, subject to applicable institutional review board (IRB) approvals, participant consent, and compliance with the Health Insurance Portability and Accountability Act (HIPAA) and other relevant privacy and confidentiality regulations

