## Supplemental Information for "Discrete cortico-hippocampal circuit states underlie mood state transitions in depression"

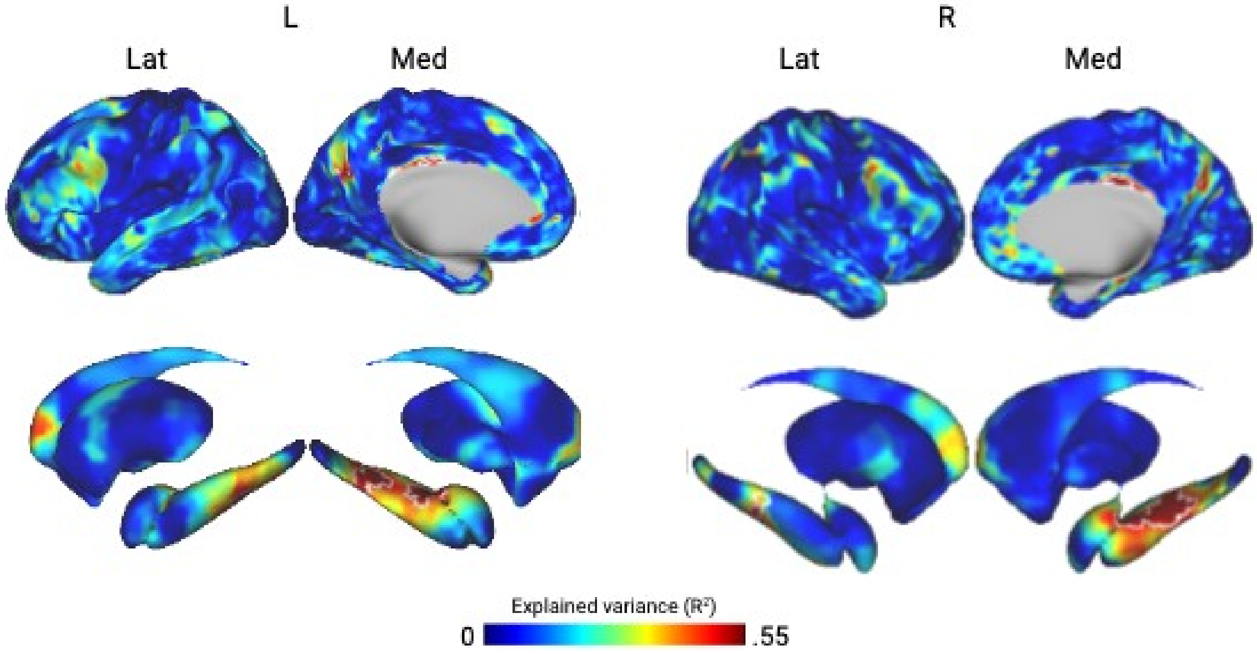


**Figure S1 | Full-surface visualization of the brain-wide dissimilarity analysis shown in Figure 1c.** Lateral (Lat) and medial (Med) cortical views of the left (L) and right (R) hemispheres, with corresponding subcortical and hippocampal surfaces, display the explained variance ($R^2$) from the regression of session-wise functional-connectivity (FC) dissimilarity against ECT-Distance in the longitudinal patient. This figure provides the complete set of surface views for the analysis summarized in Figure 1c. Permutation testing with spatial multiple-comparisons correction identified the bilateral anterior–mid hippocampus as the region whose FC changes were most strongly explained by the clinical time course; white outlines denote clusters surviving correction.

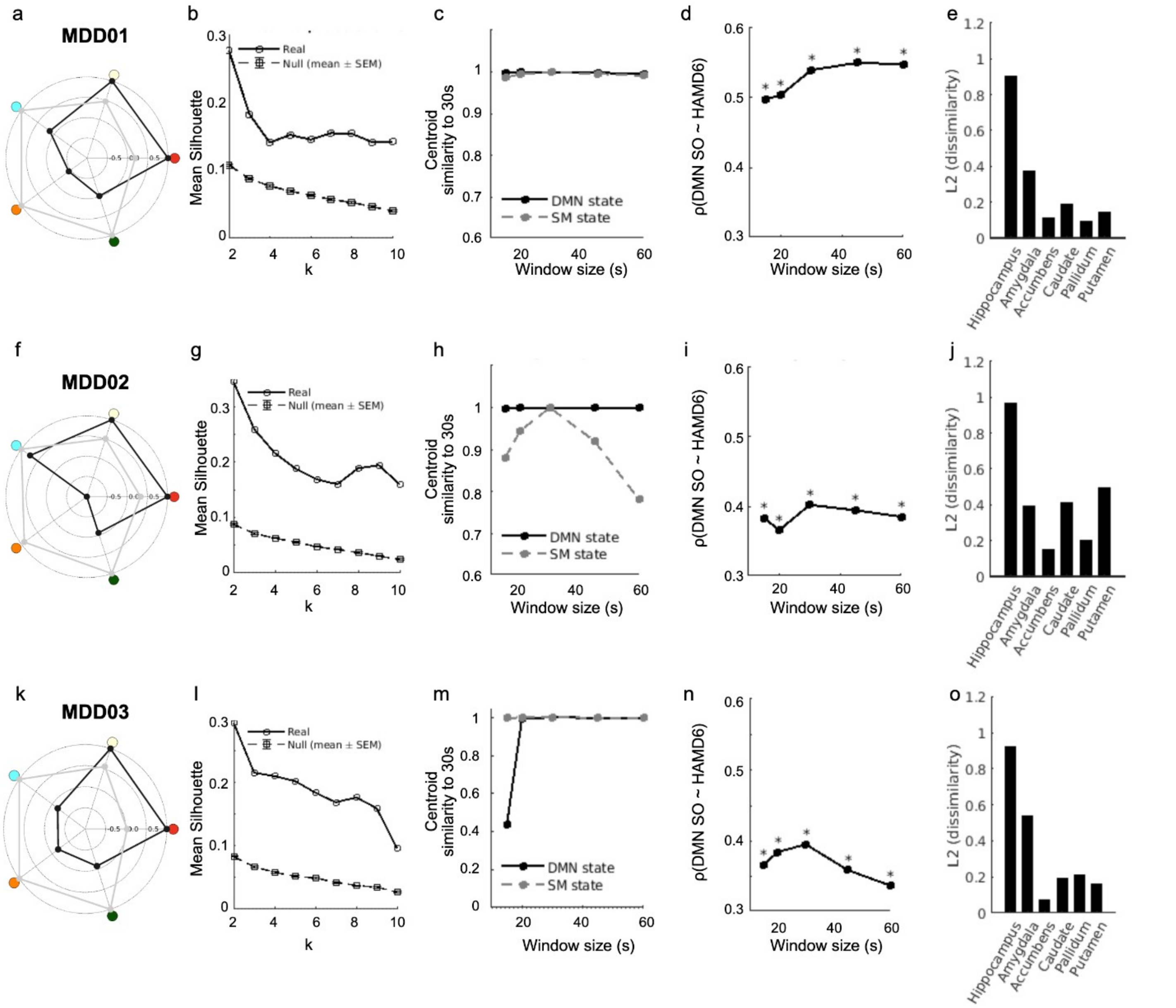


**Figure S2 | Robustness and reproducibility of hippocampal state detection across longitudinally sampled MDD patients.** (a–e) Results for patient MDD01; (f–j) for MDD02; (k–o) for MDD03. (a, f, k) Centroid radar plots display hippocampal functional connectivity (FC) with the most distinctive cortical networks for the two identified states, highlighting opposite connectivity patterns between the DMN- and SM-dominant configurations. (b, g, l) Mean silhouette values as a function of cluster number (k) demonstrate optimal separation at $k = 2$ relative to a phase-shifted null distribution (mean ± s.e.m.), confirming the presence of two recurring hippocampal connectivity states. (c, h, m) Centroid similarity across window lengths (20–60 s) relative to the canonical 30-s window shows high consistency (similarity values between 0.8 and 1.0) for both DMN- and SM-dominant states, indicating stable state definitions across temporal resolutions. The only deviation occurred in MDD03, where the SM-state centroid was less stable at the shortest (20-s) window length, reflecting reduced clustering reliability at very brief windows. (d, i, n) Correlations between DMN-state occupancy (SO) and depressive symptom severity (HAMD-6) remain significant across window lengths (Spearman ρ shown; $P < 0.05$ indicated by *). (e, j, o) Regional specificity analysis of state dissimilarity (L2 distance) across subcortical structures identifies the hippocampus as exhibiting the largest state-dependent FC change. Together, these analyses demonstrate that the two hippocampal connectivity states are robustly separable, temporally stable, and reproducible across three independent longitudinal MDD patients.

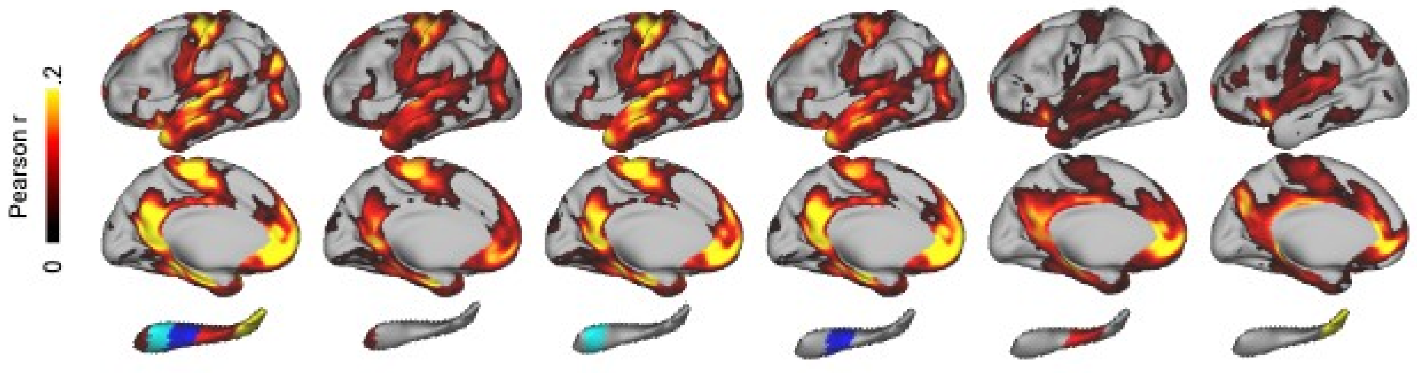


**Figure S3 | Hippocampal network connectivity along the longitudinal axis.** Functional connectivity (FC) maps show correlations (Pearson *r*) between bilateral hippocampal segments and the cortical surface, progressing from the whole hippocampus (leftmost) to anterior, mid, and posterior subdivisions (right). The anterior–mid hippocampus exhibits co-connectivity with both the default-mode (DMN) and sensorimotor (SM) networks, whereas the posterior tail primarily couples with the medial parietal network. Color scale indicates FC strength (0–0.2).

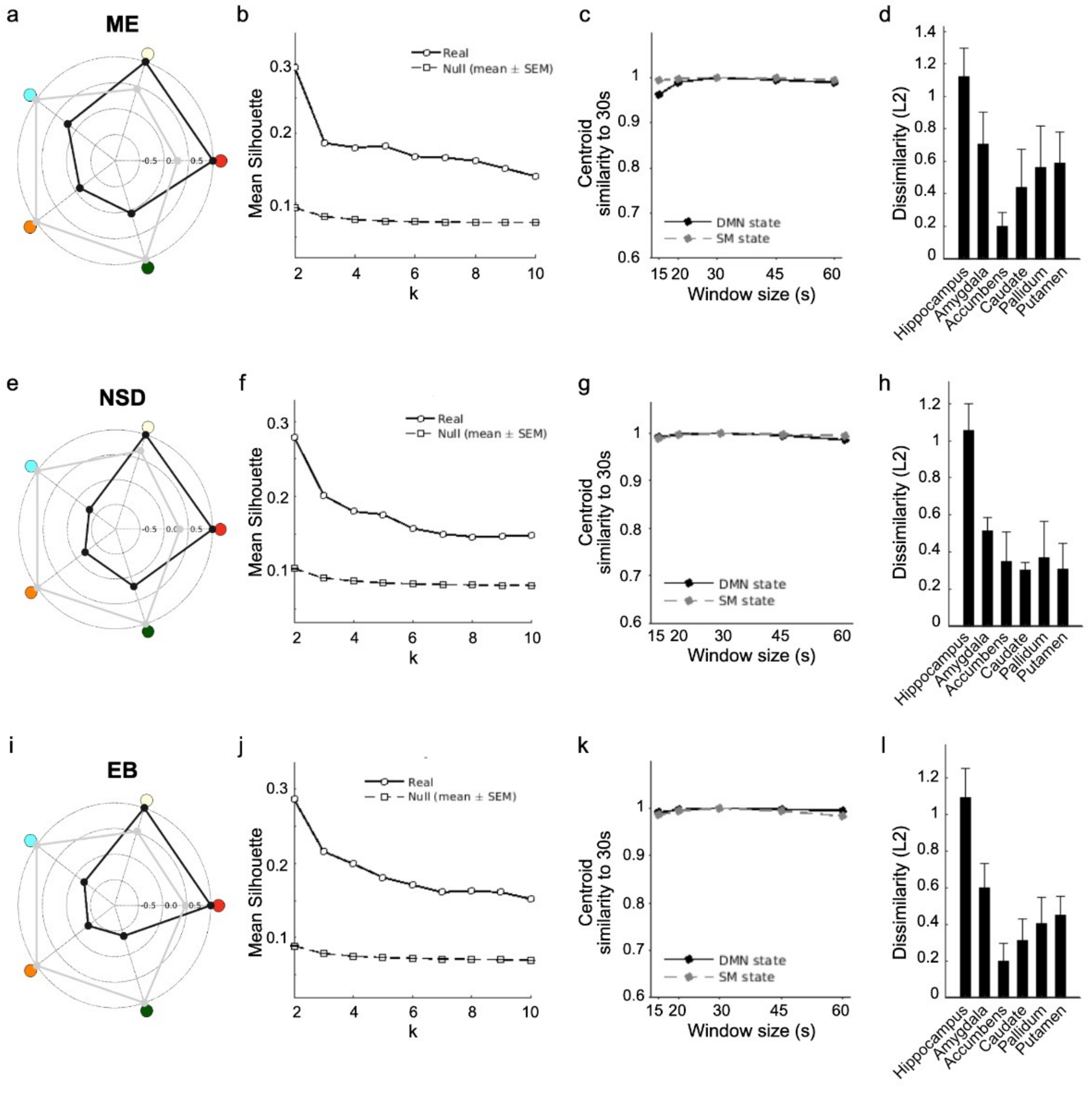


**Figure S4 | Group-level hippocampal state detection across healthy precision fMRI datasets (related to Figure 2).** (a–d) Results for the Weill Cornell multi-echo (ME) dataset; (e–h) for the 7 T Natural Scenes Dataset (NSD); (i–l) for the Ekskalibur (EB) dataset. (a, e, i) Centroid radar plots show hippocampal functional connectivity (FC) with the most distinctive cortical networks for the two identified states, highlighting opposing coupling to the default-mode (DMN) and sensorimotor (SM) systems. (b, f, j) Mean silhouette values as a function of cluster number (*k*) demonstrate an optimal solution at *k = 2* compared with a phase-shifted null distribution (mean ± s.e.m.), confirming two reproducible hippocampal connectivity states across all datasets. (c, g, k) Centroid similarity computed across window lengths (15–60 s) relative to the canonical

30-s window shows exceptionally high stability (similarity = 0.95–1.0) for both DMN- and SM-dominant states, indicating consistent state definitions across temporal scales. (d, h, l) Regional dissimilarity (L2 distance) between state-specific FC patterns across subcortical structures identifies the hippocampus as exhibiting the strongest state-dependent changes, with minimal effects in other regions. These results, derived from clustering hippocampus-to-network dynamic FC (dFC) concatenated across all sessions within each sample, confirm that the two hippocampal network states described in Figure 2 are robust, temporally stable, and reproducible across independent high-fidelity healthy fMRI cohorts.

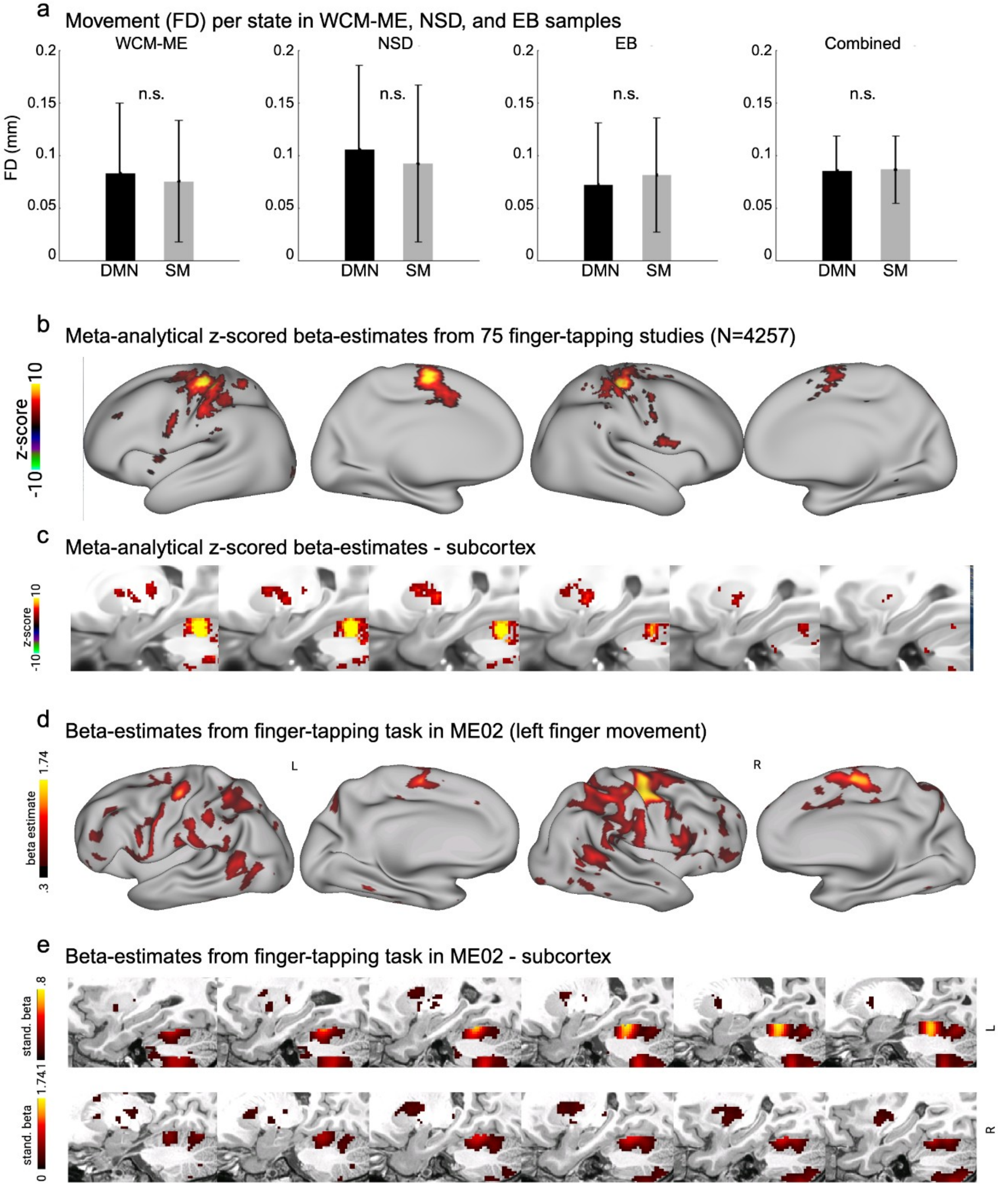


**Figure S5 | Hippocampal state dynamics are unrelated to movement, and the hippocampus does not coactivate with motor execution. (**a) Mean framewise displacement (FD) during DMN- and SM-dominant states in three densely sampled datasets (WCM-ME, NSD, and EB) and their combined sample. No significant difference in head motion was observed between states (n.s. across all comparisons). (b–c), Meta-analytic activation maps (z-scored beta-estimates) from 75 finger-tapping fMRI studies (total N = 4,257; data from Neurosynth). Cortical activation (b) is restricted to sensorimotor and premotor regions, while corresponding subcortical maps (c) show engagement of thalamic and basal-ganglia territories but no hippocampal activation. (d–e) Single-subject beta-estimates from a high-fidelity multi-echo fMRI finger-tapping task (ME02) show a similar pattern: robust activation in cortical and subcortical sensorimotor regions, absent in the hippocampus.

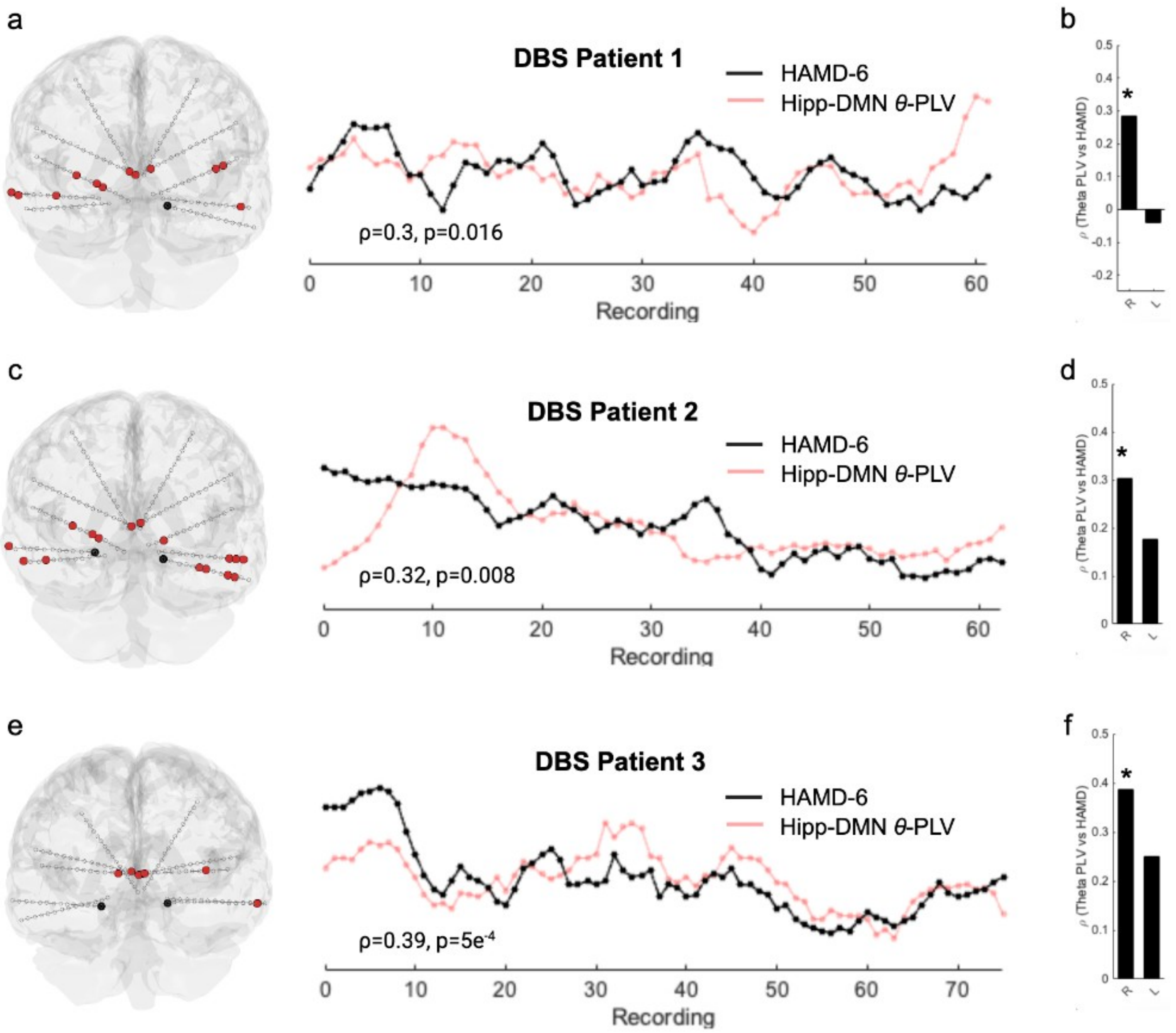


**Figure S6 | Average hippocampal–DMN theta coupling tracks depressive symptom fluctuations across intracranial EEG patients (related to Figure 4).** (a–f) Data from three patients with treatment-resistant depression implanted with bilateral depth electrodes as part of a closed-loop deep brain stimulation (DBS) trial. (a, c, e) Electrode locations show the hippocampus (black) and all cortical DMN contacts (red). Time courses depict depressive-symptom severity (HAMD-6; black) and the average theta-band (4–8 Hz) phase-locking value (PLV; red) between the right hippocampus and all ipsilateral cortical DMN contacts. Each patient shows a significant within-subject correlation between HAMD-6 and hippocampal–DMN θ-PLV (Spearman's ρ and p values indicated). (b, d, f) Correlation strength (ρ) between hippocampal–DMN θ-PLV and HAMD-6 for right (R) versus left (L) hemisphere; asterisks denote significant effects. Across all patients, correlations were significant and of similar effect size as for results shown in Fig.4. Effects were again right-lateralized. Together these results confirm the relationship between direct electrophysiological measurement of hippocampal–DMN coupling and depressive-symptom fluctuations.

| **Sample** | **Site / Scanner** | **Dx** | **Age** (years) | **N (F)** | **Voxel Size (mm)** | **TR (ms) / # ET** | **Other Parameter** | **fMRI** (minutes) | **Figure** |
|---|---|---|---|---|---|---|---|---|---|
| SIMD (*28*) | Weill Cornell Medicine / 3T Siemens Prisma | MDD /BD | 29.47 ± 8.28 | 3 (2) | 2.4 iso | 1355 / 5 | **FA:** 68° **FOV:** 216 mm **Slices:** 72 **PE:** AP **MB:** 6 **In-plane:** 2 | 1549.9 ± 367.3 | Main text Fig. 1,3, |
| WCM-TMS (*28*) | Weill Cornell Medicine / 3T Siemens Prisma | TRD | 40.89 ± 12.73 | 124 () | 2.5 iso | 1300 / 4 | **FA:** 67° **FOV:** 216 mm **Slices:** 60 **PE:** AP **MB:** 4 **In-plane:** 2 | 28.1 ± 1.3 | Main text Fig. 3 |
| WCM-ME (*26*) | Weill Cornell Medicine / 3T Siemens Prisma | HC | 33.42 ± 9.10 | 5 (0) | 2.4 iso | 1355 / 5 | **FA:** 68° **FOV:** 216 mm **Slices:** 72 **PE:** AP **MB:** 6 **In-plane:** 2 | 419.1 ± 322.9 | Main text Fig. 2, 3 |
| NSD (*87*) | University of Minnesota / 7T | HC | 26.50 ± 4.24 | 8 (6) | 1.8 iso | 1600 / 1 | **FA:** 62° **FOV:** 216 mm² **Slices:** 84 **PE:** AP **MB:** 3 **In-plane:** 0 | 115.2 ± 35.5 | Main text Fig. 2, 3 |
| EB (*70*) | Basque Center on Cognition, Brain & Language / 3T Siemens Prisma | HC | 31.4 ± 5.4 | 10 (5) | 2.4 × 2.4 × 3 | 1500 / 5 | **FA:** 70° **FOV:** 211 mm² **Slices:** 52 **PE:** AP **MB**: 4 **In-plane:** 2 | 407 ± 17.7 | Main text Fig. 2, 3 |
| HCP-PDC-ECT (*71*) | UCLA Ahmanson-Lovelace Brain Mapping Center / 3T Siemens Prisma | TRD | | 31 | 2 iso | 800 / 1 | **FA:** 52° **FOV:** 208 mm² **Slices:** 72 **PE:** AP & PA **MB**: 8 **in-plane:** 0 | 13.4 | Main text Fig. 3 |

| | | | | | | | | | |
|---|---|---|---|---|---|---|---|---|---|
| UCSF iEEG (88) | UCSF / GE 3T | TRD | 32 ± 2 | 3 (2) | 2.4 iso | 800 / 1 | **FA:** 52° **FOV:** 216 mm **Slices:** 66 **PE:** AP & PA **MB**: 6 **In-plane:** .. | 60-120 | Main text Fig.4 |

**Table S1 | Data sources.** Abbreviations: AP, anterior-posterior; BD, bipolar disorder; Dx, diagnosis; ET, echo times; F, number of female participants; FA, flip angle; FOV, field of view; HC, healthy control; iEEG, intracranial electroencephalography; iso, isotropic; MB, multiband acceleration factor; MDD, major depressive disorder; N, total number of participants; PA, posterior-anterior; PE, phase encoding direction; TR, repetition time; TRD, treatment-resistant depression.
